# Prevalence and associated risk factors for low-back pain among rehabilitation professionals in Rwanda

**DOI:** 10.1101/2025.10.13.25337817

**Authors:** Joseph Nshimiyimana, Felix Niyonkuru, Divine Girizina, Pasteur Butoya, Jackline Mupenzi Gatsinzi, Juliette Gasana, Olive Uwiduhaye, Joanitah Kemigisha, Anne Kumurenzi, Gerard Urimubenshi

## Abstract

**Background:** Rehabilitation professionals are at risk of low back pain (LBP) due to the nature of their work, which involves physical stress and awkward positioning during work. The risk factors associated with LBP can include patient transfers, repetitive tasks, lifting heavy equipment or patients.

**Objective:** This study aimed to determine the prevalence and associated risk factors for low back pain among rehabilitation professionals.

**Methods:** A cross-sectional quantitative study design employing a census sampling method was utilized in this study. A total of 274 individuals were invited to participate in this study through a Google Form questionnaire distributed via email and WhatsApp. Between January 22^nd^ and April 27^th^, 2025, 174 respondents, representing a response rate of 63.87% successfully completed the online questionnaire. These participants, all of whom work in hospital settings across Rwanda, comprised the final sample for the study. Using the Modified Nordic Questionnaire, data on different measures of LBP and risk factors were collected. The data analysis was conducted using statistical software package of IBM SPSS Statistics Version 21. Descriptive, bivariate and multivariate logistic regression analyses were conducted to identify the significant predictors of LBP.

**Results:** Among 174 participants, 78.2% reported experiencing LBP in the past year, with most cases being mild 60(34.5%) and 64(36.8%) moderate LBP. Despite this, 74.7% missed no more than three workdays annually due to LBP, and only 12.6% requested sick leave. Additionally, 26.4% said that LBP interfered with their ability to perform usual home tasks. Lifting heavy objects or patients (OR=3.152, CI=1.324-7.506, p=0.009) and physical stress (OR = 6.583, CI=2.109-20.550, p=0.009) remained statistically significant predicators, with participants engaged in these activities more likely to report LBP.

**Conclusion:** Low back pain was prevalent among rehabilitation professionals in Rwanda, predominantly with mild to moderate severity. Although LBP caused limited absenteeism, it interfered with daily activities for a large proportion. Lifting heavy objects and physical stress emerged as significant predictors of LBP, emphasising the need for workplace ergonomic interventions and strategies to reduce physical strain.

## Introduction

Low back pain (LBP) is among the most common work-related musculoskeletal disorders (WMSDs) and is currently a major social and economic concern[1]. The consequences of LBP as an occupational illness adversely affect workplaces globally and are considered one of the predominant causes of disability and medical consultations[1]. LBP is characterized by lumbar pain resulting from sprains or strains of the ligaments, tendons, or muscles in the low back, defined as discomfort located above the gluteal fold and below the 12^th^ rib [2].

While healthcare professionals in general are vulnerable to LBP due to physically demanding tasks, rehabilitation professionals are particularly at risk from factors such as patient transfers, repetitive tasks, lifting heavy equipment or patients, bending or twisting, sustaining prolonged positions during treatment [3, 4].

Throughout a standard workday, rehabilitation professionals, including physical therapists (PTs) and occupational therapists (OTs), may encounter patient care situations necessitating manual lifting or moving [5, 6]. OTs and PTs receive training in safe body mechanics and self-defense skills for the management and movement of patients inside their distinguished professional programs [2, 5]. Despite extensive training, these professionals are vulnerable to musculoskeletal issues arising from patient handling [5].

A study conducted Saudi Arabia among physical therapists [7], indicated that long-term standing (201; 44%), pushing and lifting (170; 37%), patient handling (155; 34%), and long-term sitting (129; 28%) were the work activities most adversely impacted by LBP. Furthermore, in the same study found that lifting and transferring patients were the significant predictors for developing work-related LBP. A study conducted in Zimbabwe among physical therapist included 198 physiotherapists revealed that 107 of them experienced low back discomfort, which was attributed to the risk factors for transferring and lifting clients [6].

The number of rehabilitation professionals in Rwanda is increasing, with practitioners already employed in diverse health facilities throughout the country. PTs, OTs, prosthetists, and orthotists (P&Os) provide vital rehabilitation treatments. Although these professionals play a crucial role in healthcare, they are increasingly exposed to work-related musculoskeletal risks, yet data on the prevalence and associated risk factors of such conditions in Rwanda remain scarce. The aim of this study was to determine the prevalence of low back pain and its associated risks among rehabilitation professionals in Rwanda. This study aims to bridge existing information gaps to guide targeted interventions and policies that promote the occupational health and well-being of rehabilitation professionals, hence improving the quality of rehabilitation services in Rwanda.

## Materials and methods

### Study setting

This study was conducted at three specialized hospitals, 13 teaching hospitals, and 37 provincial, district and rehabilitation institutions located in various areas of Rwanda [8, 9].

### Study design

A descriptive cross-sectional study design using the quantitative approach [10], enabled the research team to quantify the prevalence of LBP and its associated risk factors among rehabilitation professionals in Rwanda.

### Study participant’s characteristics

The study population comprised PTs, OTs, P&Os employed at specialized hospitals; teaching hospitals; provincial, district and rehabilitation institutions located in Rwanda. The participants had a minimum of one year of professional experience at the specified health facilities, with their primary responsibility being patient treatment. Therapists who were pregnant and those with previous LBP as a result from trauma or congenital anomalies were excluded. The projected study population was estimated using data from the Rwanda Health Gazette 2020 and 2022 [8, 9], which reports the number of rehabilitation professionals in the designated health facilities. Based on the records the total workforce comprises 274 professionals, including 186 PTs, 16 OTs, and 72 P&Os.

### Sampling techniques and sample size

A census sampling method was used to select the study participants[11], where all persons from the target group (274 target population) were invited to participate.

### Data collection methods and procedure

The study utilized a validated modified Nordic questionnaire [24,25], which consists of four distinct sections: (1) demographic characteristics, such as sex, age, height, weight and comorbidities, such as diabetes, obesity, hypertension, spinal problems, back surgery and arthritis; (2) professional behaviors, such as smoking habits and exercise behaviors such as walking for 15 minutes; (3) occupational-related characteristics, such as lifting heavy objects or patients, heavy workload, and sick leave due to LBP; and (4) work-related LBP, such as LBP within the last year, via the modified Nordic Questionnaire. The research team reached out to rehabilitation professionals’ associations and organizations, namely, the Rwanda Occupational Therapy Association (RWOTA), Rwanda Physiotherapy Organization (RPTO), and Rwanda Society of Prosthetists and Orthotists (RSPO), to request the contact information of registered members, including their email addresses and phone numbers. Data were collected between January 22 to April 27, 2025 via Google Form that began with an informed consent section and inclusion criteria before progressing to the main items. Throughout the four-month data collection period, the research team sent monthly reminders to participants via email or WhatsApp messages each month.

### Data analysis

The data were analysed via the Statistical Package for the Social Sciences (IBM Corp., USA) version 21. Descriptive statistics, such as frequencies, percentages, means and standard deviations, were performed for the independent variables such as age, sex, comorbidities, and walking behaviors. Bivariate analyses were conducted via the Pearson chi-square test and Fisher’s exact test to examine the relationships between LBP in the past 12 months and the sociodemographic and work-related characteristics of the rehabilitation professionals. Multivariate logistic regression was conducted to identify the significant predictors of LBP in the past 12 months, with the following independent variables: sex (male, female), age (≤35, >35), walking behaviours (< 15 minutes, ≥ 15 minutes), working experience (0-10, 11-29), lifting heavy objects or patients (yes, no), experiencing physical stress (yes, no), main daily duties (office work, treating patients), moving time in hours) (≤ 2 hours, > 2 hours), standing time in hours) (≤ 2 hours, > 2 hours), and sitting time in hours) (≤ 2 hours, > 2 hours). A P value ≤ 0.05 was considered statistically significant.

## Ethical consideration

Before starting the study, ethical approval was obtained from the Institutional Review Board (IRB) at the University of Rwanda, College of Medicine and Health Sciences (Approval Notice No. 598/CMHS IRB/2024). Permission to collect data was also requested through relevant professional organizations and associations.

The study’s aim and specific objectives were thoroughly communicated to all participants through a Google Form. Any questions or concerns submitted via the principal investigator’s email, provided within the form were addressed respectfully. To maintain confidentiality, no participant names were used in the research materials; instead, unique codes were assigned to each participant. Participation was entirely voluntary, and persons were informed that they could leave the study at any time without any consequences. The collected data were used solely for this research. All ethical practices followed the guidelines set out by the World Medical Association’s Declaration of Helsinki, which outlines ethical principles for conducting medical research with human subjects [14].

## Results

The questionnaire was administered online via Google Forms to 274 potential rehabilitation professionals through email and WhatsApp. To enhance participation, monthly reminders were issued over a four-month period. In total, 174 respondents completed the questionnaire, forming the final sample with a response rate of 63.87%. This response rate was also referenced in another study titled “Prevalence and risk factors associated with low back among health care providers in a Kuwait hospital” [15].

### Sociodemographic characteristics of the participants

The participants’ ages ranged from 23-60 years, with a mean age of 35.33 years (standard deviation 7.81). The body mass index (BMI) values ranged from 18.4 to 58.8, with a mean BMI of 24.61 and a standard deviation of 3.75. The study included a total of 174 participants, including 130 males (74.7%) and 44 females (25.3%). With respect to walking behaviours (69.0%) walked for 15 minutes or more, whereas (31.0%) walked for less than 15 minutes **(Table 1)**.

**Table 1:**
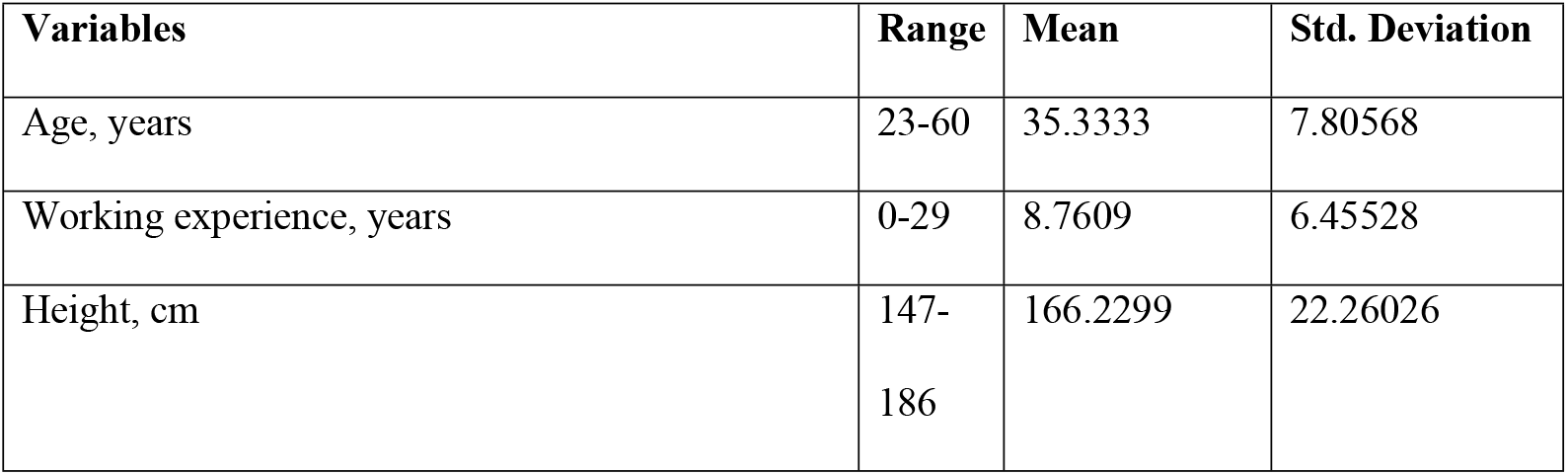
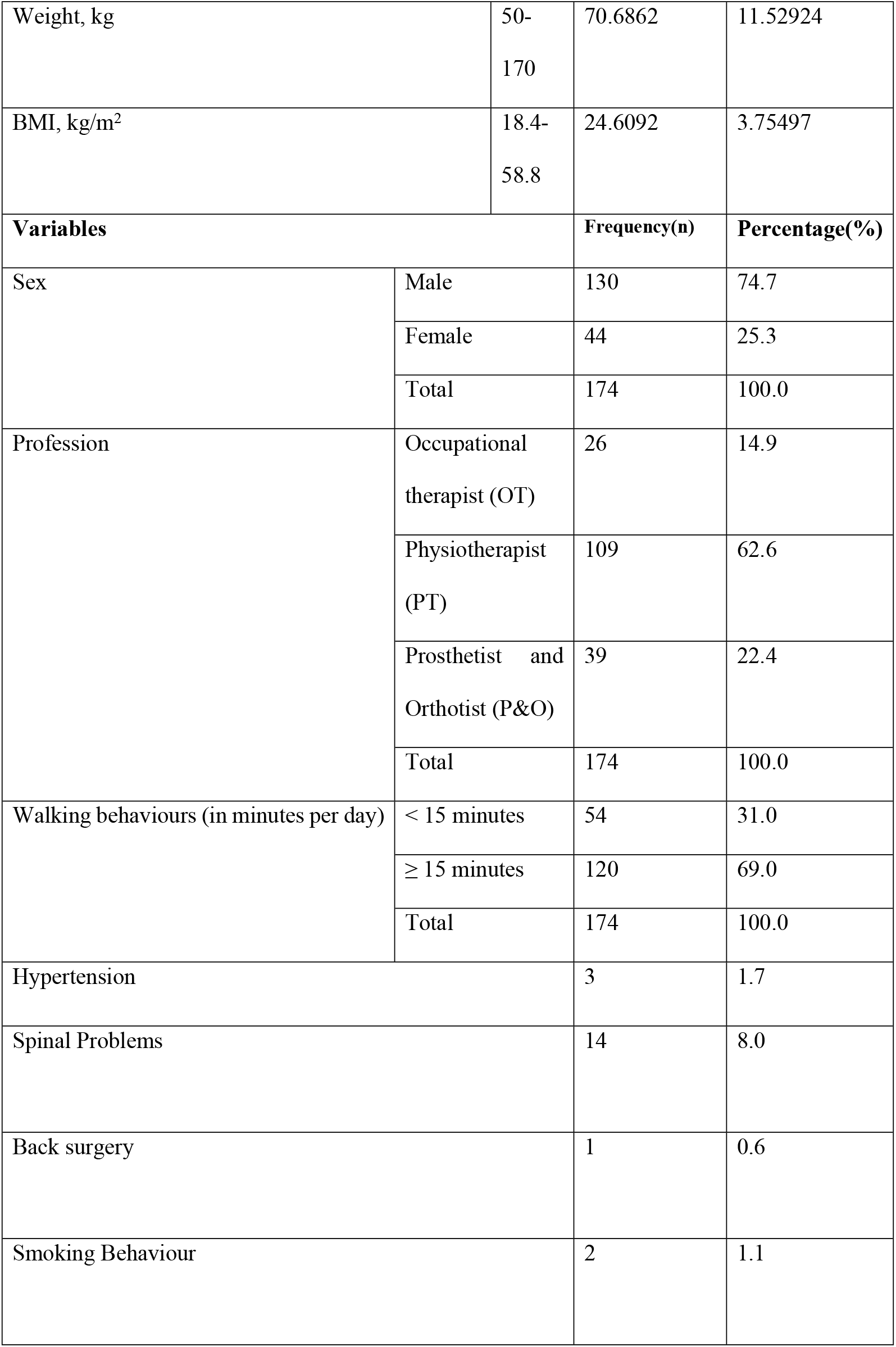
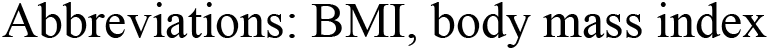
Sociodemographic characteristics of the participants.

| Variables | Range | Mean | Std. Deviation |
| --- | --- | --- | --- |
| Age, years | 23-60 | 35.3333 | 7.80568 |
| Working experience, years | 0-29 | 8.7609 | 6.45528 |
| Height, cm | 147-186 | 166.2299 | 22.26026 |

|  |  |  |  |  |
| --- | --- | --- | --- | --- |
| Weight, kg |  | 50-170 | 70.6862 | 11.52924 |
| BMI, kg/m<sup>2</sup> |  | 18.4-58.8 | 24.6092 | 3.75497 |
| **Variables** |  |  | **Frequency(n)** | **Percentage(%)** |
| Sex | Male |  | 130 | 74.7 |
|  | Female |  | 44 | 25.3 |
|  | Total |  | 174 | 100.0 |
| Profession | Occupational therapist (OT) |  | 26 | 14.9 |
|  | Physiotherapist (PT) |  | 109 | 62.6 |
|  | Prosthetist and Orthotist (P&O) |  | 39 | 22.4 |
|  | Total |  | 174 | 100.0 |
| Walking behaviours (in minutes per day) | < 15 minutes |  | 54 | 31.0 |
|  | ≥ 15 minutes |  | 120 | 69.0 |
|  | Total |  | 174 | 100.0 |
| Hypertension |  |  | 3 | 1.7 |
| Spinal Problems |  |  | 14 | 8.0 |
| Back surgery |  |  | 1 | 0.6 |
| Smoking Behaviour |  |  | 2 | 1.1 |

Abbreviations: BMI, body mass index

**Table 2:** Work-related characteristics of the participants.

| Variables |  | Frequency | Percentage |
| --- | --- | --- | --- |
| Lifting heavy objects or patients |  | 104 | 59.8 |
| Experiencing physical stress |  | 152 | 87.4 |
| Working experience, years | 0-10 | 117 | 67.2 |
|  | 11-29 | 57 | 32.8 |
| Main daily duties | Office work | 9 | 5.2 |
|  | Treating patients | 165 | 94.8 |
| Intensity of the workload | Light | 3 | 1.7 |
|  | Moderate | 118 | 67.8 |
|  | Heavy | 53 | 30.5 |
| Sitting time in hours | ≤ 2 hours | 132 | 75.9 |
|  | > 2 hours | 42 | 24.1 |
| Standing time in hours | ≤ 2 hours | 119 | 68.4 |
|  | > 2 hours | 55 | 31.6 |
| Moving time in hours | ≤ 2 hours | 130 | 74.7 |
|  | > 2 hours | 44 | 25.3 |

### Work-related characteristics of the participants

Among the 174 study participants, 104(59.8%) reported lifting heavy objects or patients and participants reported physical stress were 152(87.4%). Participant 117 (67.2%) reported 0–10 years of work experience, whereas 57 (32.8%) had 11–29 years of experience. A predominant number of participants 165(94.8%) reported that their main daily duty was patient treatment, while only 9 (5.2%) engaged in office work. In terms of workload intensity 118(67.8%) characterized their workload as moderate, and 53(30.5%) as heavy. Regarding posture-related activities, 132(75.9%) of participants reported sitting for two hours or less per day. A majority 119(68.4%) stood for no more than two hours, while 55 (31.6%) reported standing for longer periods. Most participants 130(74.7%) indicated a movement duration of two hours or less, whereas 44(25.3%) reported more than two hours.

### Prevalence of low back pain among the participants

Among the 174 participants, 136 (78.2%) reported experiencing LBP during the previous 12 months. Of those with LBP, 22 (12.6%) had requested sick leave because of the condition. Participants 46 (26.4%) indicated that LBP had interfered with their ability to perform usual household tasks at home during the past 12 months. Regarding pain severity, 60 (34.5%) participants reported mild LBP and 64 (36.8%) reported moderate LBP. Concerning work absenteeism among participants with LBP, 130 (74.7%) reported missing 0 to 3 workdays per year, and only 4(2.3%) missed between 4 to 7 days per year due to LBP **(Table 3)**.

**Table 3:**
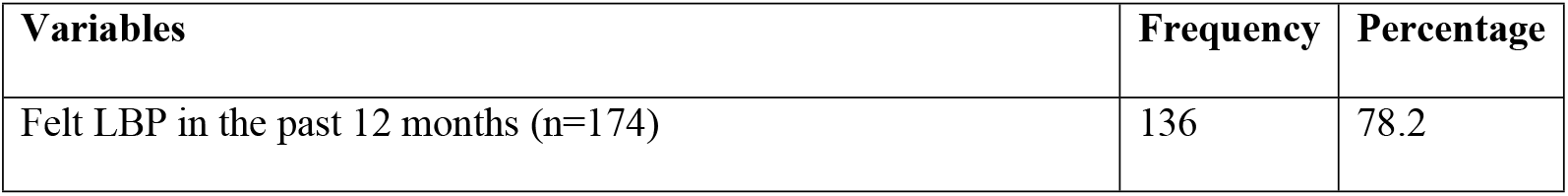
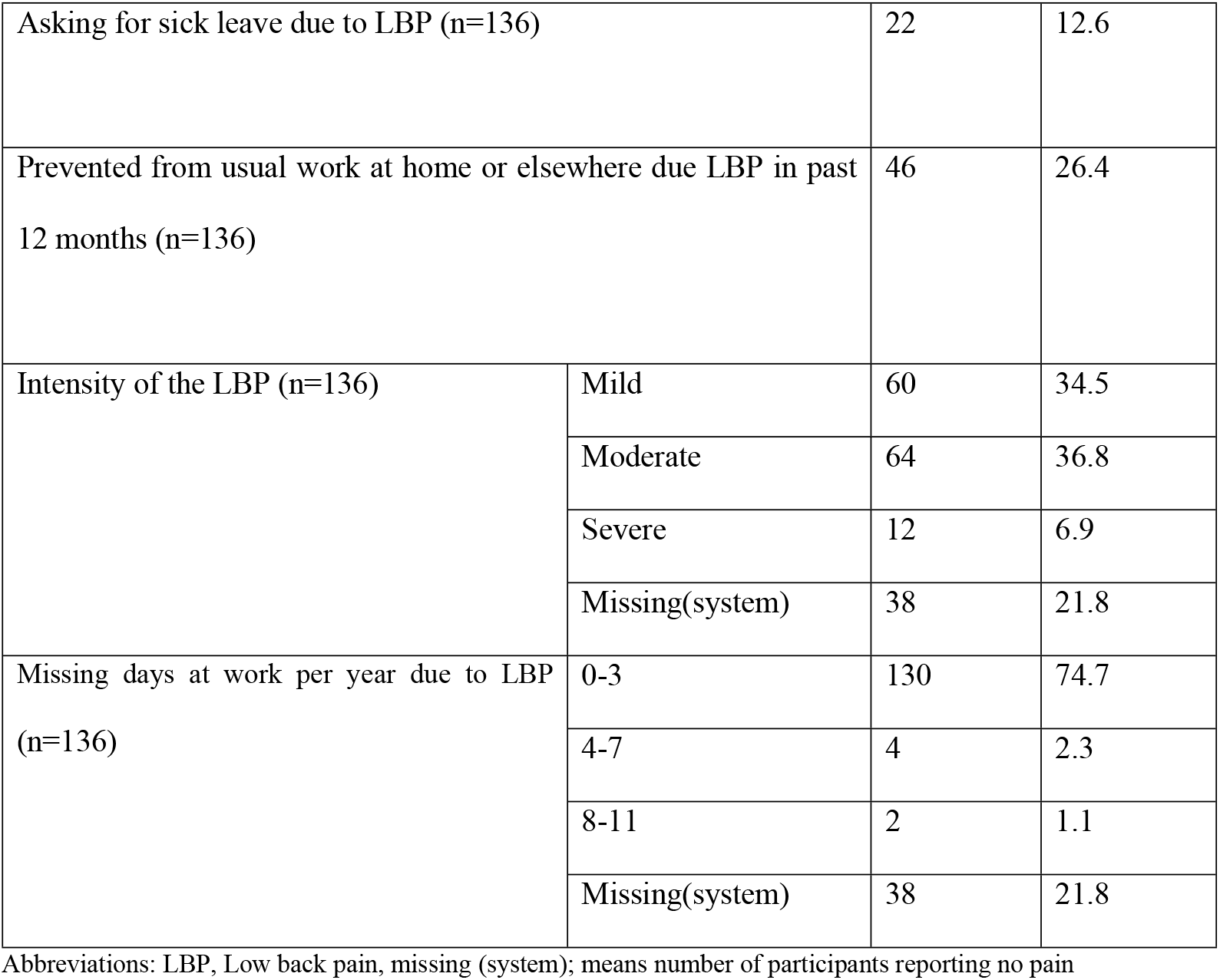
Prevalence of low back pain among the participants.

| Variables | Frequency | Percentage |
| --- | --- | --- |
| Felt LBP in the past 12 months (n=174) | 136 | 78.2 |

|  |  |  |  |
| --- | --- | --- | --- |
| Asking for sick leave due to LBP (n=136) |  | 22 | 12.6 |
| Prevented from usual work at home or elsewhere due LBP in past 12 months (n=136) |  | 46 | 26.4 |
| Intensity of the LBP (n=136) | Mild | 60 | 34.5 |
|  | Moderate | 64 | 36.8 |
|  | Severe | 12 | 6.9 |
|  | Missing(system) | 38 | 21.8 |
| Missing days at work per year due to LBP (n=136) | 0-3 | 130 | 74.7 |
|  | 4-7 | 4 | 2.3 |
|  | 8-11 | 2 | 1.1 |
|  | Missing(system) | 38 | 21.8 |

Abbreviations: LBP, Low back pain, missing (system); means number of participants reporting no pain

**Associations of low back pain (LBP) with several characteristics of the participants**

A comprehensive analysis was conducted to identify factors associated with LBP among participants, using bivariate and multivariate statistical methods. Bivariate analyses examined the relationship between LBP and each explanatory variable independently, whereas multivariate logistic regression was applied to adjust for potential confounders and determine independent predictors of LBP.

In the bivariate analyses, LBP was significantly associated with age (P=0.007), work experience (P<0.001), lifting heavy objects or patients (P<0.001), and exposure to physical stress (P<0.001). After adjustment in the multivariate model, only two variables remained statistically significant: lifting heavy objects or patients (OR = 3.152, CI: 1.324--7.506, p = 0.009) and exposure to physical stress (OR=6.583, CI: 2.109--20.550, p < 0.001) **(Table 4)**.

**Table 4:** Association of low back pain (LBP) with several characteristics of the participants.

|  |  | Bivariate Analysis |  |  | Multivariate Analysis |  |  |
| --- | --- | --- | --- | --- | --- | --- | --- |
| Variables |  | Felt LBP at any time during the past 12 months |  | P value | OR | 95% CI | P value |
|  |  | No, n (%) | Yes, n (%) |  |  |  |  |
| Sex | Male | 31 (23.8) | 99 (76.2) | 0.188 | 1.761 | 0.637-4.868 | 0.275 |
|  | Female | 7 (15.9) | 37 (84.1) |  |  |  |  |
| Age | ≤35 | 30(17.2) | 76(43.7) | **0.007** | 2.139 | 0.562-8.141 | 0.265 |
|  | >35 | 8(4.6) | 60(34.5) |  |  |  |  |
| Walking behaviors day | < 15 minutes | 10(5.7) | 44(25.3) | 0.555 | 0.806 | 0.312-2.085 | 0.657 |
|  | ≥ 15 minutes | 28(16.1) | 92(52.9) |  |  |  |  |
| Working experience, years | 0-10 | 32(18.4) | 85(48.9) | **<0.001** | 2.347 | 0.525-10.488 | 0.264 |
|  | 11-29 | 6(3.4) | 51(29.3) |  |  |  |  |
| Lifting heavy objects or patients | No | 24(13.8) | 46(26.4) | **<0.001** | **3.152** | **1.324-7.506** | **0.009** |
|  | Yes | 14(8.0) | 90(51.7) |  |  |  |  |
| Experiencing physical stress | No | 12(6.9) | 10(5.7) | **<0.001** | **6.583** | **2.109-20.550** | **<0.001** |
|  | Yes | 26(14.9) | 126(72.4) |  |  |  |  |
| Main daily duties | Office Work | 3(1.7) | 6(3.4) | 0.309 | 2.691 | 0.438-16.534 | 0.285 |
|  | Treating patient | 35(20.1) | 130(74.7) |  |  |  |  |
| Sitting time in hours | ≤ 2 hours | 29(16.7) | 103(59.2) | >0.999 | 1.013 | 0.347-2.958 | 0.981 |
|  | > 2 hours | 9(5.2) | 33(19.0) |  |  |  |  |
|  | ≤ 2 hours | 29(16.7) | 90(51.7) | 0.324 | 1.812 | 0.690-4.759 | 0.228 |
| Standing time in<br>hours | > 2 hours | 9(5.2) | 46(26.4) |  |  |  |  |
| Moving time in<br>hours | ≤ 2 hours | 29(16.7) | 101(58.0) | >0.999 | 0.849 | 0.305-2.361 | 0.754 |
|  | > 2 hours | 9(5.2) | 35(20.1) |  |  |  |  |

Abbreviations: CI, confidence interval; LBP, low back pain; OR, odds ratio; n, frequency; %, percentage

## Discussion

This paper highlights that LBP is a prevalent occupational health concern among rehabilitation professionals in Rwanda. This research provides foundational information to delineate the prevalence and associated risk factors among rehabilitation practitioners working in hospital settings in Rwanda.

The prevalence of LBP identified in this study among rehabilitation specialists in hospital settings in Rwanda is broadly consistent with findings from other studies. The prevalence of LBP in our sample was 78.2%, closely aligning with 78.1% reported by Takrouni et al. [16] among physical therapists in Saudi Arabia. Similarly, Cınar-Medeni et al. [17] reported a 12 months prevalence of LBP as 75% among healthcare professionals while Landry et al. [15] found 70.9% among healthcare providers in a Kuwaiti hospital. Within Rwanda, a study conducted at the Military Hospital reported a prevalence of LBP of 78% among nurses [18], further supporting the notion that LBP is a prevalent occupational health issue across different healthcare disciplines.

Our findings also highlight the functional impact of LBP on rehabilitation professionals. Twenty-two (12.6%) participants taking sick leave because back pain, highlighting its contribution to work-related absenteeism. Pain intensity varied, with 64 (36.8%) participants reporting moderate symptoms and 12 (6.9%) describing severe pain, suggesting the need for targeted preventive and therapeutic strategies for those at greater risk.

Despite the high prevalence of LBP, most participants 130 (74.7%) reported missing only 0-3 workdays annually due to LBP. This pattern may indicate that many professionals continue working despite discomfort, potentially because occupational pressures, limited access to treatment, or effective personal coping mechanisms. These findings highlight the importance of early detection, workplace ergonomics, and timely to prevent or relieve the impact of LBP on productivity and overall quality of life.

Age was also found to influence the occurrence of LBP in our sample. Among professionals aged≤35 years, 43.7% reported LBP, compared with 34.5% among those >35 years and this difference was statistically significant (p=0.007). This finding is consistent with the systematic review by Rezaei et al. [19], which reported that age maybe related to LBP among healthcare professionals; although the association was weak. Other studies, including that by Landry et al. [15] and Ovayolu et al. [20], found no significant relationship between age and LBP among healthcare practitioners. One explanation for these inconsistencies may be the age distribution of study samples. Although the cited study revealed no correlation between age and LBP, a significant majority of participants (68.0% in Group A and 73.0% in Group B) were aged between 20 and 40 years. The restricted age distribution may have constrained the capacity to identify significant disparities among age groups. The underrepresentation of older professionals (e.g., those over 40 years or 50 years) undermines the statistical ability to evaluate age-related patterns.

Conversely, the study demonstrating significance likely had a wider age range or possessed a more equitable sample, facilitating clearer comparisons between younger and older professionals. Furthermore, a study conducted by revealed no correlation between age and low back pain among healthcare workers.

These findings indicate that age is not strongly associated with low back pain among healthcare professionals, despite certain specific instances in which age may be associated with this demographic. This tendency may indicate that cumulative physical stresses and musculoskeletal deterioration are linked to aging, underscoring the necessity for age-specific preventive measures. This study revealed that young professionals with between 0 and 10 years of work experience have the highest prevalence of low back pain.

This study found that persons with 0–10 years of experience reported a markedly greater incidence of low back pain (48.9%) than their more seasoned peers did. The p value of <0.001 strongly indicates that young professionals with ten years or less of work experience are more likely to develop low back discomfort. This aligns with the study undertaken by Ezzatvar et al. [21], which revealed that reduced job experience is connected with musculoskeletal discomfort among physical therapists. In a manner similar to the study titled “Work-Related Factors Associated with Low Back Pain Among Nurse Professionals in East and West Wollega Zones, Western Ethiopia, 2017: A Cross-Sectional Study” conducted by Mekonnen [22], our findings diverge, revealing that low back pain is correlated with nurses with limited professional experience, specifically those with ≤ 5 years. This may result from younger professionals undertaking more physically demanding activities, lacking ergonomic training, or being less adept at controlling occupational strain. It emphasizes the importance of early intervention, encompassing ergonomic education and workload management, for professionals at the beginning of their careers. Given these findings, specific methods should be formulated to address LBP among rehabilitation professionals.

This study revealed two variables as statistically significant predictors of low back pain among rehabilitation professionals: lifting heavy objects or patients and experiencing physical stress, as determined by both bivariate and multivariate analyses. Rehabilitation professionals involved in lifting duties are more than three times as likely to report low back pain (LBP), with an odds ratio (OR) of 3.152 and a multivariate p value of 0.009. This aligns with the findings of Karahan et al. [23], who reported a significant correlation between low back pain among healthcare professionals and the lifting of heavy objects, as determined via multivariate logistic regression analysis. A study by McCrory et al. [24] showed results analogous to ours, indicating that lifting heavy objects or gadgets among physical rehabilitation therapists is correlated with low back pain. These findings indicate that physical exertion from manual handling significantly contributes to musculoskeletal discomfort.

This study discovered that physical stress had an odds ratio of 6.583 and a highly significant p value of <0.001 in both analyses. This signifies that those undergoing physical stress are almost six times more likely to obtain LBP, rendering it the most significant predictor in the sample. In alignment with the study titled “Epidemiology of work-related low back pain among rehabilitation professionals in Saudi Arabia” by Abolfotouh et al. [5], physical stress was shown to be significantly correlated with low back pain among rehabilitation professionals. Similar to other healthcare professionals, nurses see a significant correlation between physical stress and low back pain [25].

The duration of sitting, standing, or moving during the day does not seem to affect the probability of developing LBP, as evidenced by elevated p values and odds ratios near 1. This implies that the duration of posture may be less significant than the quality or intensity of physical activity in this type of study. In light of these findings, it is advisable for hospital settings to emphasize ergonomic training and safe lifting techniques, especially for those engaged in patient handling.

Furthermore, managing physical stress via workload regulation, wellness initiatives, and stress alleviation techniques may considerably diminish the prevalence of low back pain (LBP). Although duration in different postures does not seem to be a significant risk factor, emphasis should be placed on the manner in which tasks are executed rather than the length of time they are maintained. These insights can guide targeted interventions and policy modifications to enhance occupational health and alleviate the burden of low back pain among healthcare workers.

This study examined the prevalence status and related risk factors of LBP among rehabilitation specialists employed in hospital settings in Rwanda. However, it was unable to examine the preventive techniques utilized by rehabilitation specialists to avert low back discomfort. Consequently, the study to be undertaken in this context should incorporate preventive interventions.

## Conclusion

LBP is common among rehabilitation professionals in hospital settings in Rwanda. The low back pain prevented the rehabilitation experts from engaging in their customary duties for some time each year, prompting many to request leave owing to discomfort, with many experiencing moderate pain intensity. This occurrence is associated with substantial lifting of objects or patients and physical strain as primary predictors of low back pain among rehabilitation specialists. Rehabilitation professionals should receive training in workplace ergonomics, proper body positioning, techniques for simplifying tasks, strategies for reducing workload, and the importance of sharing responsibilities when tasks exceed individual physical capacity.

## Data Availability

All data produced in the present study are available upon reasonable request to the authors

## Acknowledgement

The research team sincerely acknowledges the valuable time and effort contributed by the participants in completing the online survey.

## Authors contributions

NJ, NF, and GD developed the research proposal and involved in data collection.UG supervised this research project and facilitated the strong data analysis. GJ, BP, OU and KJ were involved in data collection and cleaning. NJ, UG, GJ AK and KJ contributed in manuscript preparation and editing.

